# *Mycobacterium avium* complex (MAC) genomics and transmission in a London hospital

**DOI:** 10.1101/2022.01.07.22268791

**Authors:** Andries J van Tonder, Huw C Ellis, Colin P Churchward, Kartik Kumar, Newara Ramadan, Susan Benson, Julian Parkhill, Miriam F Moffatt, Michael R Loebinger, William OC Cookson

## Abstract

Non-tuberculous mycobacteria (NTM) are ubiquitous environmental microorganisms and opportunistic pathogens in individuals with pre-existing lung conditions such as cystic fibrosis (CF) and non-CF bronchiectasis (BX). Whilst recent studies of *Mycobacterium abscessus* have identified transmission within single CF centres as well as nationally and globally, transmission of other NTM species is less well studied. We sequenced 996 Mycobacterium avium complex (MAC) isolates from CF and non-CF patients at the Royal Brompton Hospital (RBH), London. Genomic analysis was used to analyse local transmission. Epidemiological links were identified from patient records. These and previously published genomes were used to characterise global population structures. Analysis of the three predominant MAC species identified putative transmission clusters that contained patients with CF, BX and other lung conditions, although few epidemiological links could be identified. For *M. avium*, lineages were largely limited to single countries, whilst for *M. chimaera*, global transmission clusters previously associated with heater cooler units (HCUs) were found. However, the immediate ancestor of the lineage causing the major HCU-associated outbreak was a lineage already circulating in patients with pre-existing lung conditions. CF and non-CF patients shared transmission chains even in the presence of CF patient-focussed hospital control measures, although the lack of epidemiological links suggested that most transmission is indirect and may involve environmental intermediates or else asymptomatic carriage in the wider population. The major HCU-associated *M. chimaera* lineage being derived from an already circulating lineage, suggests that HCUs, while being responsible for a major global transmission event, are not the sole vector nor the ultimate source of this wider patient-infecting lineage. Future studies should include sampling of environmental reservoirs and potential asymptomatic carriers.

**Author summary:** Whilst recent studies in *Mycobacterium abscessus* have identified transmission within single CF centres as well as nationally and globally, the transmission dynamics between CF and non-CF patients has not yet been comprehensively examined in the *Mycobacterium avium* complex (MAC). We believe this is the first study to use a well-sampled longitudinal isolate dataset, that includes both CF and non-CF patients from a single hospital setting, to investigate transmission of MAC species. We identified transmission clusters in the three predominant MAC species circulating in the hospital and showed that these included both CF and non-CF patients. We then incorporated isolates from previous studies to examine the global population structure of MAC species and showed that for *M. avium* there were UK-specific lineages circulating amongst patients, whilst for *M. chimaera* we could identify global lineages associated with HCUs. For the first time, we also show that the predominant HCU-associate lineage is likely derived from already circulating lineages associated with patients with respiratory diseases. Our study shows the value of integrating whole genome sequencing with epidemiological data to perform high-resolution molecular analyses to characterise MAC populations and identify transmission clusters. Knowledge of putative transmission networks can improve responses to outbreaks and inform targeted infection control and clinical practice.

## Introduction

Non-tuberculous mycobacteria (NTM) are ubiquitous environmental microorganisms found in soil and water and are considered opportunistic pathogens in humans. Individuals with pre-existing genetic or acquired lung diseases such as cystic fibrosis (CF), non-CF bronchiectasis (BX) and chronic obstructive pulmonary disease (COPD) are more prone to NTM disease although individuals with no known immune dysfunction can also present with NTM infections (1–3). Symptoms of NTM pulmonary disease are variable but most patients will develop a chronic cough and other symptoms may include fatigue, sputum production, chest pain, breathlessness, fever and weight loss (1). Globally, disease due to NTM infections is increasing in prevalence. For example, the estimated prevalence of NTM disease in the United States of America (USA) rose from 2.4 cases/100,000 in the early 1980s to 15.2 cases/100,000 in 2013 (4), whilst in the United Kingdom (UK) the prevalence rose from 0.9 cases/100,000 to 2.9 cases/100,000 between 1995 and 2006 (5). NTM infections may be progressive and treatment requires prolonged multi-drug therapy (6) and is often unsuccessful due to an absence of antimicrobial agents with low toxicity and effective *in vivo* activity against NTM species (1).

A number of NTM species including *Mycobacterium abscessus* and members of the *Mycobacterium avium* complex (MAC), notably *M. avium* and *M. intracellulare*, have emerged as major respiratory pathogens in the past three decades (7–9). Another member of the MAC, *M. chimaera*, has also been implicated in numerous global infections associated with cardiothoracic surgery with the source of infections linked to heater-cooler units (HCUs) contaminated during their manufacture (10–12).

Until recently the prevailing hypothesis was that infections caused by NTM were due to independent acquisitions from environmental sources such as soil, contaminated drinking water distribution systems and household plumbing. Recent studies of *M. abscessus* in CF patients have however identified indirect patient-patient transmission within a single CF centre as well as the presence of globally circulating clones of *M. abscessus* amongst CF patients worldwide (13–17). An observational study of *M. abscessus*, across England revealed that these dominant clones are also found in patients with other chronic respiratory diseases. The study however was unable to identify epidemiological links for most closely-related isolates, suggesting environmental acquisition may be involved (18). Another recent study of *M. abscessus* has demonstrated that transmission networks may involve both people with CF and those without, and that these transmission networks are global. It is therefore likely that transmission is complex, involving multiple patient cadres as well as environmental intermediates (19). In the special case of *M. chimaera*, the high level of genetic similarity between sequenced *M. chimaera* isolates collected from patients, HCUs and the factory of origin suggested a point source contamination during manufacture causing global distribution followed by localised transmission (12).

To date, little work has been done to examine whether similar patterns of transmission in the MAC are occurring between patients with CF, BX or other chronic respiratory diseases. Using a large collection of longitudinal isolates collected from patients attending the Royal Brompton Hospital (RBH) in London, the aims of this study were to characterise the population structure of MAC; to identify potential transmission chains involving patients with CF and other non-CF lung conditions; and to place the RBH isolates in a global context using previously published genomes.

## Methods

### Consent/ethics

The study (access to patients’ clinical data) was approved by the NHS Health Research Authority (HRA) and Health and Care Research Wales (HCRW) (REC reference 21/HRA/2554).

### Data collection

Clinical data pertaining to patients from whom NTM cultures were isolated were collected from electronic health records at the RBH. Data included patients’ sex, age at the time of first positive NTM culture, height, weight, lung function test results, comorbidities, medication history and date of death (where applicable). Anonymization was undertaken by removing personal data, including patients’ hospital numbers, prior to analysis.

### Sample collection

Isolates were collected from patients attending the respiratory inpatient and outpatient clinics of the RBH between January 2013 and April 2016. The RBH routinely archive all mycobacterial isolates cultured from their patients and this archive was used without selection as the basis for the study.

### Culturing, DNA extraction and sequencing

Sputum samples were grown in BBL MGIT media (BD) in a Bactec MGIT 960 (BD) until the system indicated Mycobacterial growth. Species confirmation was performed using the HAIN GenoType System. Confirmed MAC cultures were regrown from bead stock cultures in BBL MGIT media (BD) in a Bactec MGIT 960 (BD) until the system indicated growth. In the absence of growth, DNA was extracted from the bead stock. DNA extractions were performed as previously described (https://dx.doi.org/10.17504/protocols.io.bf28jqhw). A total of 1189 DNA extracts were sequenced by the core pipeline teams at the Wellcome Sanger Institute. The Illumina Hiseq X10 platform was used to generate 2 × 150 bp paired-end reads. Raw sequencing reads were deposited at the European Nucleotide Archive under project PRJEB21813. All accessions used in this project are listed in S1 Table.

### Sequence QC, mapping and phylogenetics

Basic quality control metrics for the raw sequence data were generated using FastQC v0.11.9 (20). Sequence reads with similarity to *Mycobacterium* species were identified using Kraken v0.10.6 (21) and Bracken v1.0 (22). Samples with < 70% reads mapping to a *Mycobacterium* species were excluded from further analyses (n = 116). Seven isolates not belonging to the MAC (*M. abscessus, M. chelonae, M. simiae)* were removed from the dataset. Sequence reads for each species were trimmed using Trimmomatic v0.33 (23) and mapped to appropriate references (Table A in S1 Data) using BWA mem v0.7.17 (minimum and maximum insert sizes of 50 bp and 1000 bp respectively) (24). Single nucleotide polymorphisms (SNPs) were called using SAMtools v1.2 mpileup and BCFtools v1.2 (minimum base call quality of 50 and minimum root squared mapping quality of 30) as previously described (25). Samples with reads that mapped to < 80% of the reference were excluded (n = 70). Variant sites were extracted from the resulting alignments using snp-sites v2.5.1 (26). Whole species maximum likelihood phylogenetic trees were built using IQ-tree v1.6.5 accounting for constant sites (-fconst; determined using snp-sites -C) with the built-in model testing (-m MFP) to determine the best phylogenetic model and 1000 ultrafast bootstraps (-bb 1000) (27).

For higher-resolution phylogenies within fastBAPS lineages, recombinant regions were identified and removed from alignments using GUBBINS (28) and new phylogenetic trees were constructed as described above. Pairwise SNP distances were calculated for all pairs of isolates using pairsnp (29).

### Global collections

To provide context for each the isolates sequenced for each species in this study, datasets consisting of published sequenced isolates were assembled (S2 Table) (12,30–49). Sequence data were downloaded from the European Nucleotide Archive (ENA) and trimmed; Sample QC, mapping and phylogenetic tree construction were performed as detailed above. Only the first isolate from each patient was included from the RBH isolates for each species/subspecies.

### Genome assemblies

A previously published pipeline was used to produce annotated assemblies (50). Briefly, sequence reads were assembled with spades v 3.10.10 (51) and assemblies were improved by first scaffolding the assembled contigs using SSPACE v2.0 (52) and filling the sequence gaps with GapFiller v1.11 (53).

### Transmission and epidemiological linkage

Genomic lineages were identified using fastBAPS (54) and new alignments were created for lineages ≥ ten isolates by aligning sequence reads for included isolates against the assembly that had the smallest number of contigs (using the method described above). In order to calculate a pairwise SNP threshold to determine putative transmission clusters within each genomic lineage, pairwise SNP distances for all isolates for each species in the RBH datasets were calculated. Using a previously described method (55), the transmission threshold for each species, regardless of lineage, was calculated by taking the 95^th^ percentile of the maximum within-patient isolate pairwise SNP distances for all patients and adding twice the number of mutations expected to occur in a six month period. To account for excess within-patient diversity observed in the *M. chimaera* FB1 and *M. avium* subsp. *avium* FB14 lineages (Fig A in S1 Data), pairwise SNP distances greater than 25 and 50 (assumed to result from infection with multiple lineages) were removed respectively before the above calculations were performed. Based on these results, the R library iGRAPH (56,57) and pairwise SNP thresholds of 16 (*M. intracellulare* and *M. avium* subsp. *hominissuis*), 30 (*M. chimaera*) and 58 SNPs (*M. avium* subsp. *avium*) were used to calculate putative transmission clusters in each genomic lineage. Finally, in order to identify possible epidemiological links between patients infected within the same transmission clusters, hospital stay records were examined for epidemiological contacts. The latter were defined as patients attending the same ward on the same day up to one year prior to the collection of the first sequenced isolate.

## Results

### Patient demographics

The median age and BMI of the 354 patients included in the study was 56 years (range 5 - 93) and 22.5 (range 13.4 – 43.4) respectively. One hundred and seventy-four patients (49.2%) were male and 38/354 (10.7%) were smokers. There were 147/354 (41.5%) patients with BX, 87/354 (24.6%) with CF, 53/354 (15.0%) with COPD, 32/354 (9.0%) with asthma, 19/354 (5.4%) with allergic bronchopulmonary aspergillosis (ABPA), 17/354 (4.8%) with interstitial lung disease (ILD) and 7/354 (2.0%) with other underlying respiratory conditions such as pleural thickening or sarcoidosis. Patients with no other respiratory disease or predisposition to NTM infections accounted for 3.4% (12/354) of the cohort. During the study period, 55/354 (15.5%) patients were on antibiotic treatment regimes. For 20 patients (5.6%) clinical data were unavailable.

### Species distribution

A total of 996 isolates from 354 patients were successfully sequenced and the nine MAC species identified are detailed in Table 1. The three predominant species amongst the sequenced isolates were *M. avium (M. avium* subsp. *avium* and *M. avium* subsp. *hominissuis), M. chimaera* and *M. intracellulare*. Together these accounted for 926/996 (93.0%) of the MAC isolates sequenced. Most patients were infected with only a single species during the collection period. However, 45 of the 354 patients (12.7%) were infected with two or more species (Fig 1A). In this group of patients, most of the isolates collected were typically from a single species, with other species observed more infrequently (Fig 1B). Subsequent analyses in this study will focus on the three predominant species in the dataset: *M. intracellulare, M. avium (M. avium* subsp. *avium* and *M. avium* subsp. *hominissuis)*, and *M. chimaera*.

**Table 1.** *Mycobacterium avium* complex species identified in the Royal Brompton Hospital Collection.

| Species | Number of genomes |
| --- | --- |
| *M. avium* | 405 |
| *M. chimaera* | 359 |
| *M. intracellulare* | 162 |
| *Mycobacterium* sp. MOTT36Y | 39 |
| *M. paraintracellulare* | 15 |
| *M. yongonense* | 10 |
| *M. marseillense* | 4 |
| *M. colombiense* | 1 |
| *Mycobacterium* sp. QIA-37 | 1 |

**Fig 1.**
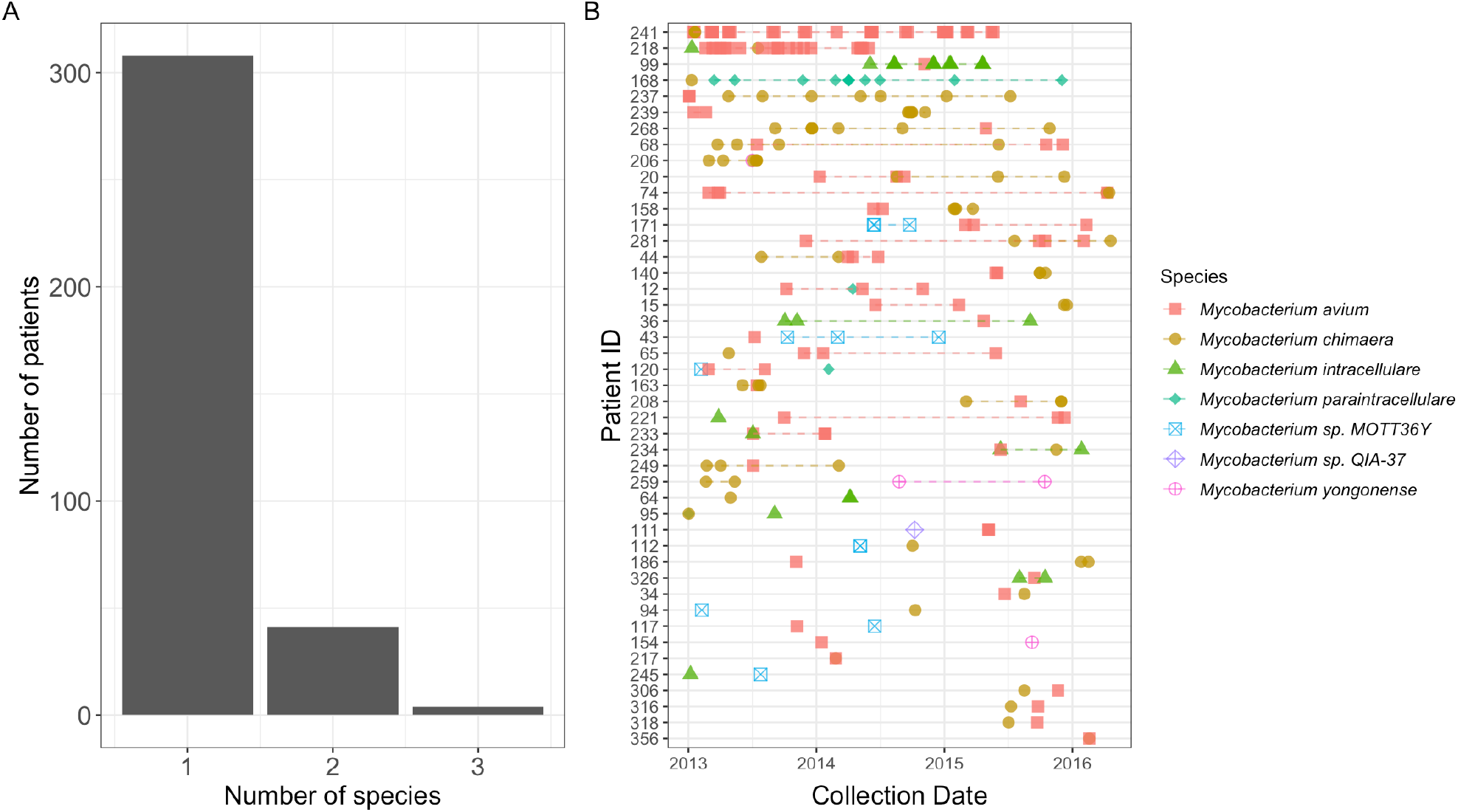
Patients infected by more than one *Mycobacterium avium* Complex (MAC) species. A) Histogram showing the number of *Mycobacterium avium* complex species identified in each patient; B) Timeline of patients infected by more than one *Mycobacterium avium* Complex (MAC) species. Each point represents an isolate and is coloured according to the species identified. Isolates from the species in a patient are linked together. Patients with one isolate or multiple isolates from a single species were excluded.

### M. intracellulare

A total of 162 genomes from 37 patients were identified as *M. intracellulare* (Fig 2A). Eleven of the patients had CF, seventeen had BX, with the remaining seven having other lung conditions (COPD n = 3; ILD n = 3; asthma n = 1; congenital pulmonary airway malformation [CPAM] n = 1) and disease metadata were missing for two patients. Genomic clustering with fastBAPs identified nine lineages with three of these having more than ten genomes (Fig 2A). Following remapping to local references for the three largest fastBAPS lineages, three putative transmission clusters were identified with the largest, Mi_FB3_1, composed of 16 patients (Fig 2B; Table B in S1 Data). Of these 16 patients, eight had BX, seven CF and one ILD. The range of the number of isolates collected per patient was between one and 32. Four of the sixteen patients were also infected with other species or lineages during the sampling period with Mi_FB3_1 only being detected in 1/22 isolates collected for patient 218 (Fig 2B). During the time period that the sequenced isolates were collected, four of the patients (85, 95, 97 and 99), were being treated with antibiotics and three of the four patients were treated successfully. No epidemiological links were identified between patients in the year prior to isolate collection. Comparison with 77 previously published *M. intracellulare* isolates showed that the most closely related contextual isolate was collected in the UK in 2015/2016 [32] and formed part of a clade containing isolates from the Mi_FB3_1 cluster (Fig 2C). The remainder of the RBH isolates were evenly distributed through the global phylogenetic tree.

**Fig 2:**
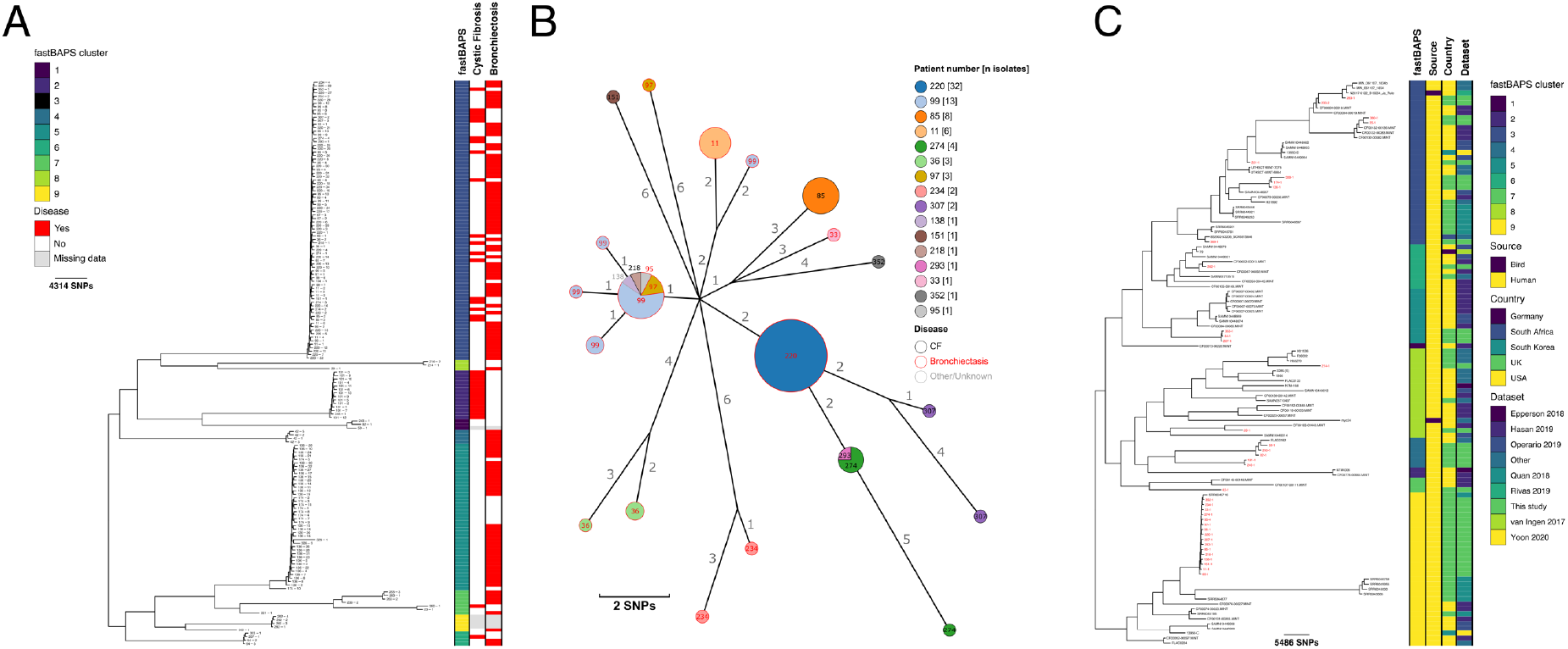
Population structure and transmission of *Mycobacterium intracellulare*. A) Midpoint-rooted maximum likelihood phylogenetic tree for *Mycobacterium intracellulare* isolates collected at the Royal Brompton Hospital (n = 162). The taxa are clustered according to their sequence similarities. The lengths of the branches are scaled in nucleotide substitutions per site. The disease status of the patients (that isolates were collected from) for cystic fibrosis and non-cystic fibrosis bronchiectasis are represented by red bars for Yes and white bars for No with missing patient data shown by gray bars. Scale bars are shown in SNPs per site.; B) *Mycobacterium intracellulare* Mi_FB3_1 transmission cluster. Each node represents an isolate or isolates identical (0 SNPs) to each other and the size of the node is proportional to the number of identical isolates. Nodes are coloured and labelled by patient number. The outer ring of each node is coloured according to the disease status of the patient (cystic fibrosis = black, non-cystic fibrosis bronchiectasis = red, grey = other lung condition). The edges represent the pairwise SNP distance between the isolate(s); C) Midpoint-rooted maximum likelihood phylogenetic tree for global *Mycobacterium intracellulare* isolates (n = 114). The taxa are clustered according to their sequence similarities. The lengths of the branches are scaled in nucleotide substitutions per site. fastBAPS lineage, source of isolate, country of collection and study are shown as datastrips to the right of the phylogenies. Isolates from the Royal Brompton Hospital are highlighted in red. Scale bars are shown in SNPs per site.

### M. avium

A total of 405 sequenced isolates collected from 176 patients were identified as *M. avium*. Of the patients infected with *M. avium*, 44 had CF and 81 BX. Sequence data for all *M. avium* isolates was mapped to a single reference (*M. avium* subsp. *avium* 104; NC008595.1) and a phylogenetic tree constructed with a previously published *M. avium* subsp. *paratuberculosis* isolate (DRR263663) chosen as an outgroup (Fig B in S1 Data). The structure of this phylogeny showed that there were two major clades which corresponded to the two subspecies *M. avium* subsp. *avium* (n = 207; MAA) and *M. avium* subsp. *hominissuis* (n = 198; MAH). Isolates for each of the two subspecies were next analysed separately. A small number of patients were infected with both subspecies (n = 6). No examples of the other two *M. avium* subspecies, *paratuberculosis* and *silvaticum*, were identified in this study.

### M. avium avium (MAA)

Of the 76 patients infected with MAA, 15 had CF, 39 had BX and 16 had other lung conditions (COPD n = 10; ILD n = 3; ABPA n = 3; asthma n = 6) whilst three patients, including one smoker, had no pre-existing respiratory disease (metadata were unavailable for three patients; Fig 3A). Clustering of the 207 MAA genomes identified 17 fastBAPS lineages (Fig 3A). Remapping of the four largest fastBAPS lineages allowed the identification of seven putative transmission clusters comprising between two and ten patients (Table C in S1 Data) with the largest cluster of ten isolates comprised of patients with CF (n = 2), BX (n = 6) and asthma or COPD (n =2; Fig 3B).

**Fig 3.**
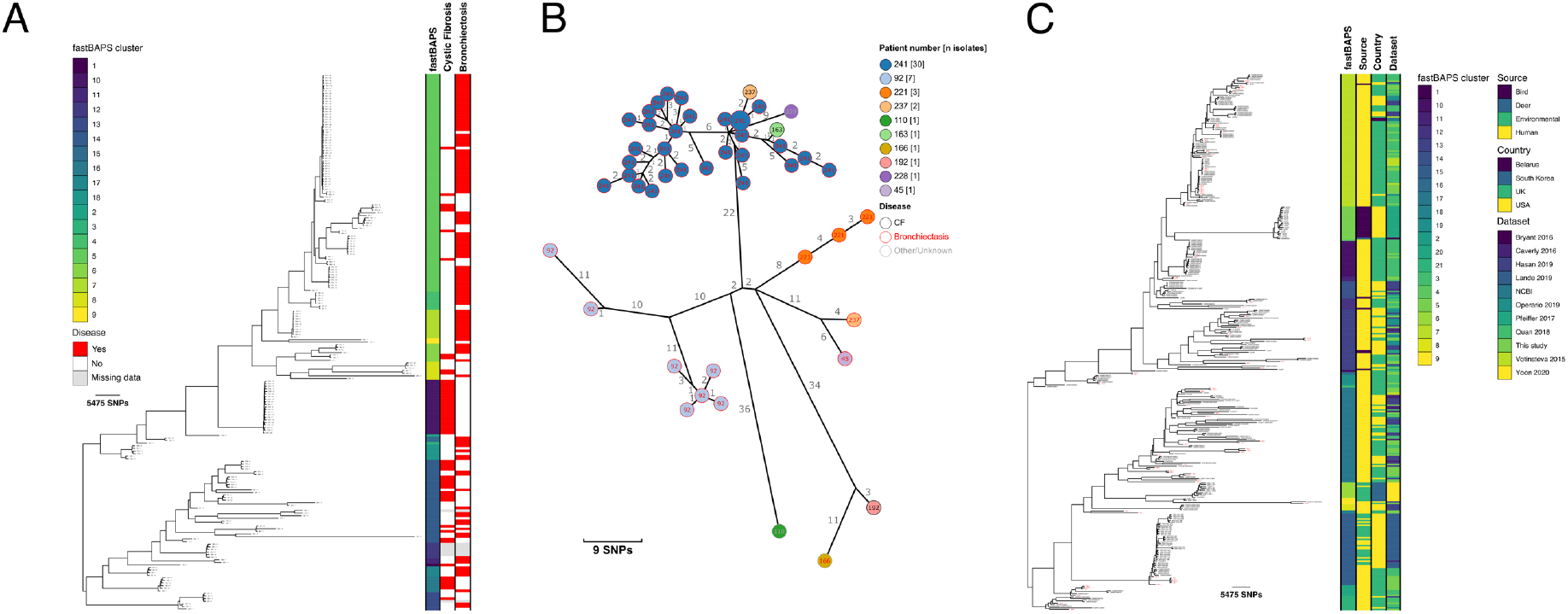
Population structure and transmission of *Mycobacterium avium* subsp. *avium*. A) Midpoint-rooted maximum likelihood phylogenetic tree for *Mycobacterium avium* subsp. *avium* isolates collected at the Royal Brompton Hospital (n = 207). The taxa are clustered according to their sequence similarities. The lengths of the branches are scaled in nucleotide substitutions per site. The disease status of the patients (that isolates were collected from) for cystic fibrosis and non-cystic fibrosis bronchiectasis are represented by red bars for Yes and white bars for No with missing patient data shown by gray bars. Scale bars are shown in SNPs per site; B) *Mycobacterium avium* subsp. *avium* MAA_FB5_1 transmission cluster. Each node represents an isolate or isolates identical (0 SNPs) to each other and the size of the node is proportional to the number of identical isolates. Nodes are coloured and labelled by patient number. The outer ring of each node is coloured according to the disease status of the patient (cystic fibrosis = black, non-cystic fibrosis bronchiectasis = red, grey = other lung condition). The edges represent the pairwise SNP distance between the isolate(s); C) Midpoint-rooted maximum likelihood phylogenetic tree for global *Mycobacterium avium* subsp. *avium* isolates (n = 344). The taxa are clustered according to their sequence similarities. The lengths of the branches are scaled in nucleotide substitutions per site. fastBAPS lineage, source of isolate, country of collection and study are shown as datastrips to the right of the phylogenies. Isolates from the Royal Brompton Hospital are highlighted in red. Scale bars are shown in SNPs per site.

A total of 21 fastBAPS lineages were defined in the global phylogeny (Fig 3C). Examination of the distribution of pairwise SNP distances for each fastBAPS lineage containing at least ten genomes revealed that there were two lineages containing RBH isolates with lower median pairwise SNP distances (FB10 and FB15; Fig 3C and Fig CB in S1 Data). The 23 genomes, including two RBH genomes, in FB10 were collected in the UK between 2013 and 2016, with most coming from the National Mycobacterial Reference Service. The latter characterises mycobacterial cultures from across the Midlands and North of England (Fig 5A) (37). If the SNP threshold used to calculate putative transmission clusters circulating in the RBH was applied, then one isolate from the RBH (40-1) formed part of a cluster comprised of isolates from Quan *et al*. (37). The majority (35/46) of the isolates forming FB15 were collected in the USA with most from a study investigating *M. avium* in the community and household water in Philadelphia (Fig 5B) (33). The pairwise distances between two of the RBH isolates and other UK isolates and isolates from the USA in FB15 were within the threshold used to define transmission, suggesting potential cross-Atlantic transmission (Fig DA in S1 Data).

**Fig 4.**
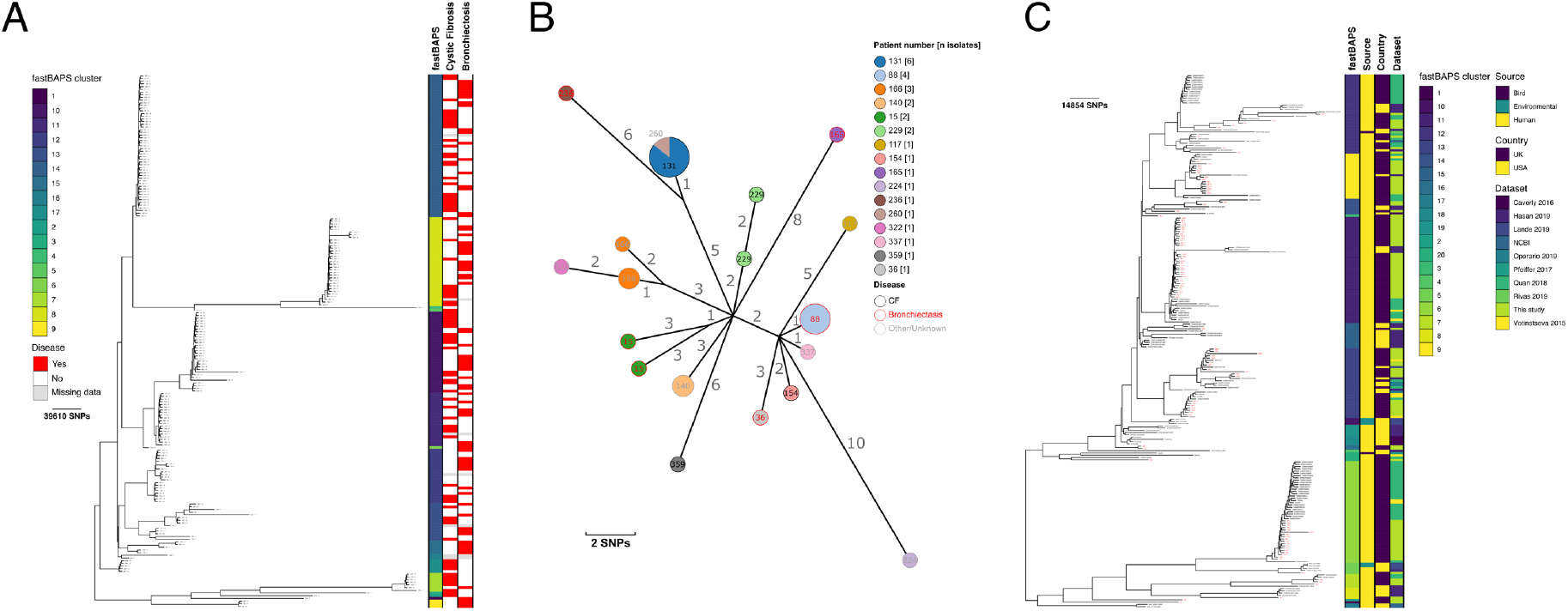
Population structure and transmission of *Mycobacterium avium* subsp. *hominissuis*. A) Midpoint-rooted maximum likelihood phylogenetic tree for *Mycobacterium avium* subsp. *Hominissuis* isolates collected at the Royal Brompton Hospital (n = 198). The taxa are clustered according to their sequence similarities. The lengths of the branches are scaled in nucleotide substitutions per site. The disease status of the patients (that isolates were collected from) for cystic fibrosis and non-cystic fibrosis bronchiectasis are represented by red bars for Yes and white bars for No with missing patient data shown by gray bars. Scale bars are shown in SNPs per site; B) *Mycobacterium avium* subsp. *hominissuis* MAH_FB8_1 transmission cluster. Each node represents an isolate or isolates identical (0 SNPs) to each other and the size of the node is proportional to the number of identical isolates. Nodes are coloured and labelled by patient number. The outer ring of each node is coloured according to the disease status of the patient (cystic fibrosis = black, non-cystic fibrosis bronchiectasis = red, grey = other lung condition). The edges represent the pairwise SNP distance between the isolate(s); C) Midpoint-rooted maximum likelihood phylogenetic tree for global *Mycobacterium avium* subsp. *hominissuis* isolates (n = 236). The taxa are clustered according to their sequence similarities. The lengths of the branches are scaled in nucleotide substitutions per site. fastBAPS lineage, source of isolate, country of collection and study are shown as datastrips to the right of the phylogenies. Isolates from the Royal Brompton Hospital are highlighted in red. Scale bars are shown in SNPs per site.

**Fig 5:**
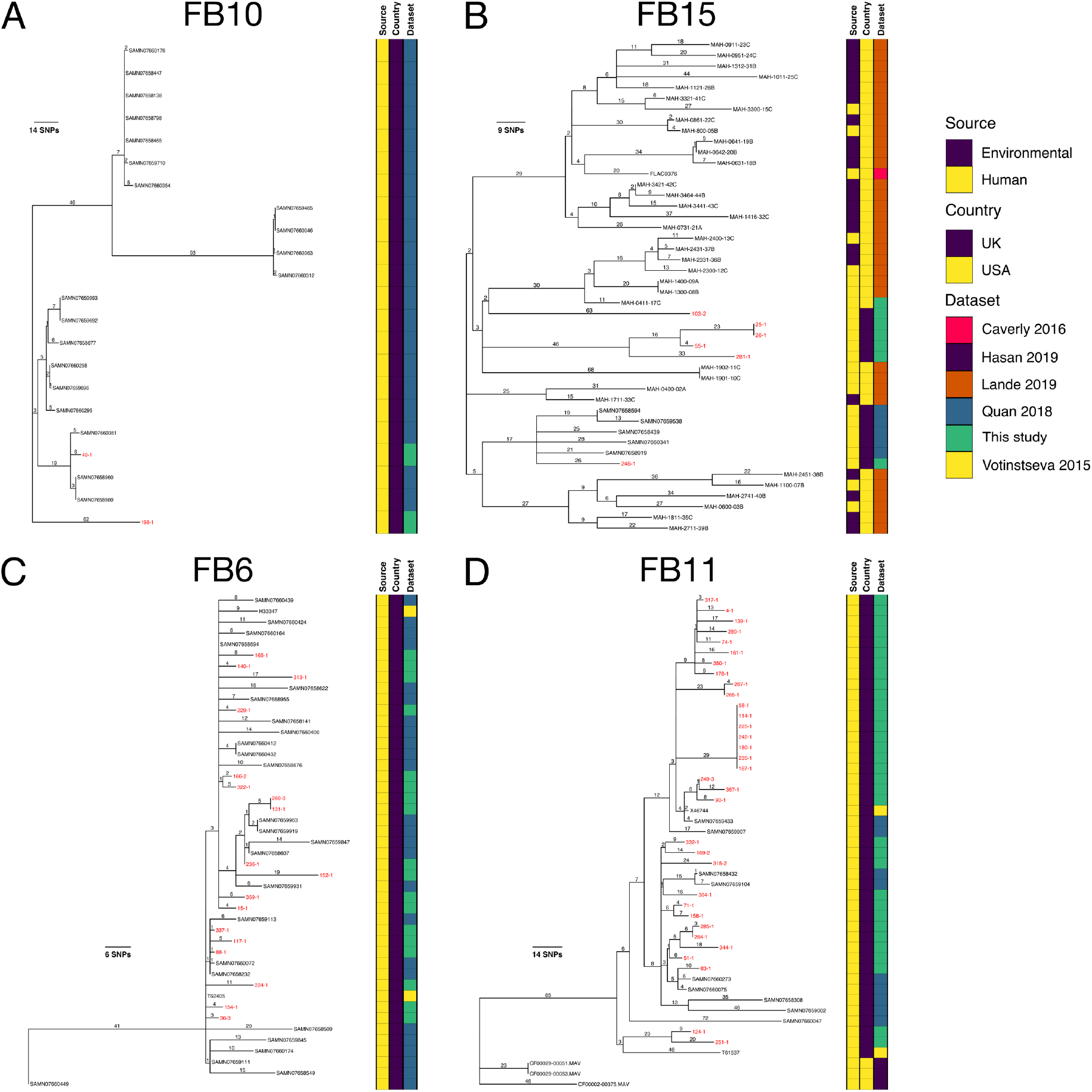
*Mycobacterium avium* global fastBAPS phylogenies. Midpoint-rooted maximum likelihood phylogenetic trees for A) *Mycobacterium avium* subsp. *avium* FB10; B) *Mycobacterium avium* subsp. *avium* FB15; C) *Mycobacterium avium* subsp. *hominissuis* FB6 and D) *Mycobacterium avium* subsp. *hominissuis* FB11. The taxa are clustered according to their sequence similarities. The lengths of the branches are scaled in nucleotide substitutions per site. The source of isolate, country of collection and study are shown as datastrips to the right of the phylogenies. Isolates from the Royal Brompton Hospital are highlighted in red. Branch lengths are shown in SNPs per site.

### M. avium hominissuis (MAH)

A total of 198 genomes from 106 patients were characterised as MAH (Fig 4A). Thirty-three patients had CF, 43 BX and 21 had other lung conditions (COPD n = 14; ILD n = 1; ABPA n = 5; sarcoidosis n = 1; asthma n = 11). Four patients had no pre-existing respiratory disease. Disease metadata were unavailable for the remaining five patients. Genomic clustering identified 17 lineages and seven putative transmission clusters containing between two and 16 patients (Table D in S1 Data). The largest cluster of 16 isolates is shown in Figure 4B and comprised patients with CF (n = 4), BX (n = 7), COPD (n = 3) and a single patient with no pre-existing lung condition (metadata were unavailable for one patient). Two pairs of patients in each of the clusters, MAH_FB8_1 and MAH_FB14_9, were found to have epidemiological links.

Of the 20 defined fastBAPS lineages in the global collection, FB6 and FB11 had lower median pairwise SNP distances and contained RBH isolates (Fig 4C and Fig CC in S1 Data). Both lineages were composed completely or near-completely of isolates collected in the UK (Fig 5C and Fig 5D). The star-like structure of the FB6 phylogeny is suggestive of a point source outbreak and applying a transmission SNP threshold of 16 SNPs showed that 41/45 of the isolates in FB6 would have formed a transmission cluster that included both isolates from this study and isolates from other UK studies (Fig DB in S1 Data) (35,37). Two small transmission clusters, containing RBH and other UK isolates, were also identified in FB11.

### M. chimaera

There were 359 sequenced isolates collected from 155 patients identified as *M. chimaera*; 37 of the patients had CF, 60 had BX whilst 50 patients had other lung conditions (COPD n = 24; ILD n =10; ABPA n = 11; pleural thickening with enfolded lung n = 1; lung non-mucinous adenocarcinoma n = 1; Wegener’s granulomatosis n = 1; *M. tuberculosis* in lymph nodes during same endoscopy procedure n = 1; primary ciliary dyskinesia n = 1; asthma n = 17). There were five patients with no pre-existing respiratory disease and metadata were unavailable for a further nine patients. A single predominant lineage, Mc_FB1, corresponding to the previously characterised Group 1 accounted for 332/359 (92.5%) of the sequenced isolates with the majority of the remaining isolates (25/359; 7.0%) forming a second main lineage Mc_FB2 corresponding to Group 2 (Fig 6A). Thirteen putative transmission clusters containing two to 106 patients were identified (Table E in S1 Data). Twelve potential epidemiological links between 15 different patients were found in transmission cluster Mc_FB3_1, the largest transmission cluster of 106 patients (Fig 6B; Table F in S1 Data). Of the patients in this cluster 43 had BX, 24 CF, 13 COPD, 14 asthma, seven ILD, two ABPA, one pleural thickening with enfolded lung, one lung non-mucinous adenocarcinoma and one primary ciliary dyskinesia. Three patients had no pre-existing lung condition and disease status was missing for five patients. Fifteen patients underwent antibiotic treatment during the study period.

**Fig 6.**
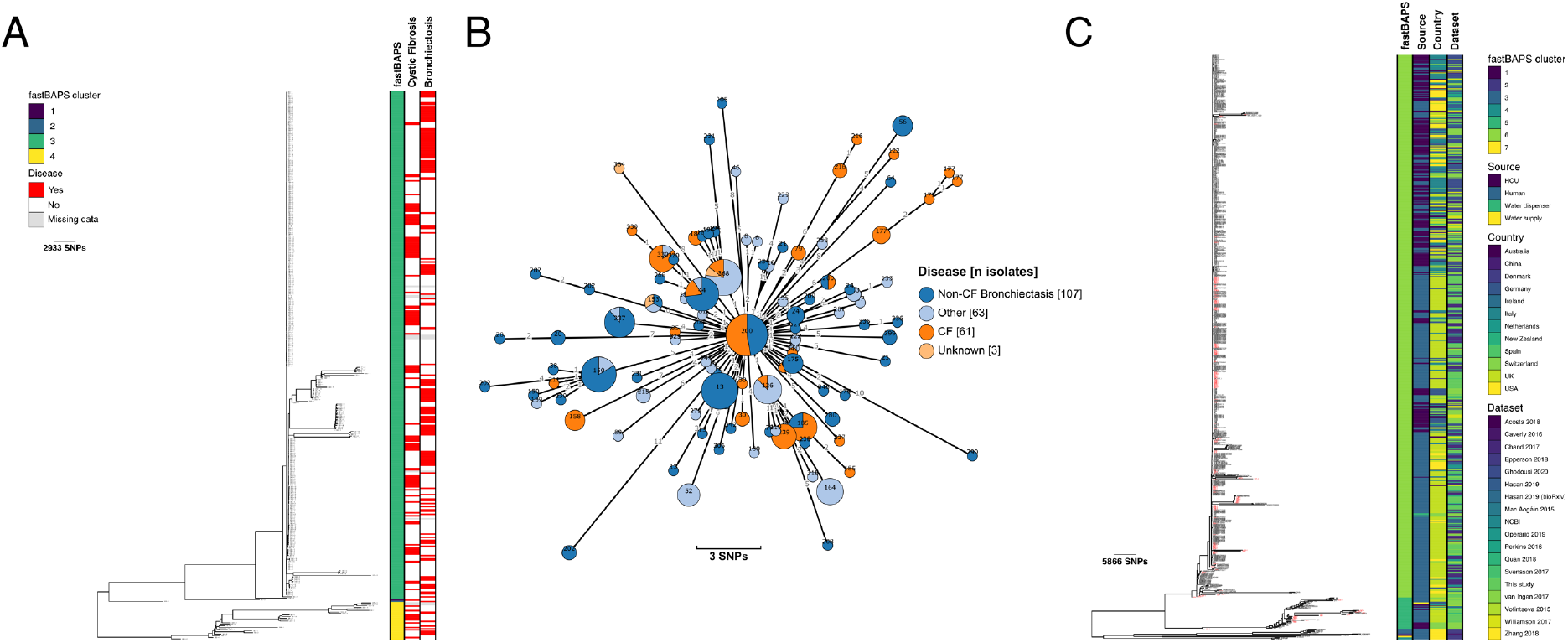
Population structure and transmission of *Mycobacterium chimaera*. A) Midpoint-rooted maximum likelihood phylogenetic tree for *Mycobacterium chimaera* isolates collected at the Royal Brompton Hospital (n = 359). The taxa are clustered according to their sequence similarities. The lengths of the branches are scaled in nucleotide substitutions per site. The disease status of the patients (that isolates were collected from) for cystic fibrosis and non-cystic fibrosis bronchiectasis are represented by red bars for Yes and white bars for No with missing patient data shown by gray bars. Scale bars are shown in SNPs per site; B) *Mycobacterium chimaera* Mc_FB3_1 transmission cluster. Each node represents an isolate or isolates identical (0 SNPs) to each other and the size of the node is proportional to the number of identical isolates. Nodes are coloured according to patient disease status and labelled by patient number. The edges represent the pairwise SNP distance between the isolate(s); C) Rooted maximum likelihood phylogenetic tree for global *Mycobacterium chimaera* isolates (n = 826). The taxa are clustered according to their sequence similarities. The lengths of the branches are scaled in nucleotide substitutions per site. fastBAPS lineage, source of isolate, country of collection and study are shown as datastrips to the right of the phylogenies. Isolates from the Royal Brompton Hospital are highlighted in red. Scale bars are shown in SNPs per site.

The global phylogenetic tree containing 155 isolates from RBH and 671 previously published isolates was topologically similar to the tree built using only the isolates from RBH with a single major clade with low genetic diversity present (FB6; Fig 6C). Isolates from the two clusters with more than ten representatives (FB5 and FB6) were remapped to local references. Recombination was removed and new phylogenetic trees constructed (Fig 7A [FB5] and Fig 7B [FB6]). Most isolates in FB5 were collected from patients with respiratory conditions or else patients with infections following cardiac surgeries. There was, however, a distinct sub-clade rooting within this diversity containing only isolates from HCUs or hospital water supplies (Fig 7B). Within FB6 a more deeply rooting diverse clade contained mainly human isolates and the larger shallower clade rooting within this contained most of the HCU isolates interspersed with human isolates (Fig 7B and Fig 7C). Due to the large number of isolates assigned to FB6 (n = 765), further genomic lineage assignment was performed using fastBAPS to identify five sub-lineages within FB6 (Fig 7B and Fig 7C). New mapping was performed for the three lineages with more than ten isolates (FB6_FB1, FB6_FB2 and FB6_FB4; Fig 8A-8C;). Two of these lineages, FB6_FB2 and FB6_FB4, were completely or nearly completely composed of patient isolates (Fig 8B and Fig 8C). Most of the isolates collected from HCUs or water supplies made up the bulk of FB6_FB1, though human isolates were distributed throughout the tree (Fig 8A). Putative transmission clusters were calculated for all five fastBAPS lineages described above used a pairwise SNP threshold of 30 SNPs (Table G in S1 Data). Transmission clusters containing RBH isolates were found in each of the lineages with the largest, found in FB6_FB1, comprising 258 HCU isolates, 230 patient isolates and a single isolate from a water supply (Table G in S1 Data).

**Fig 7.**
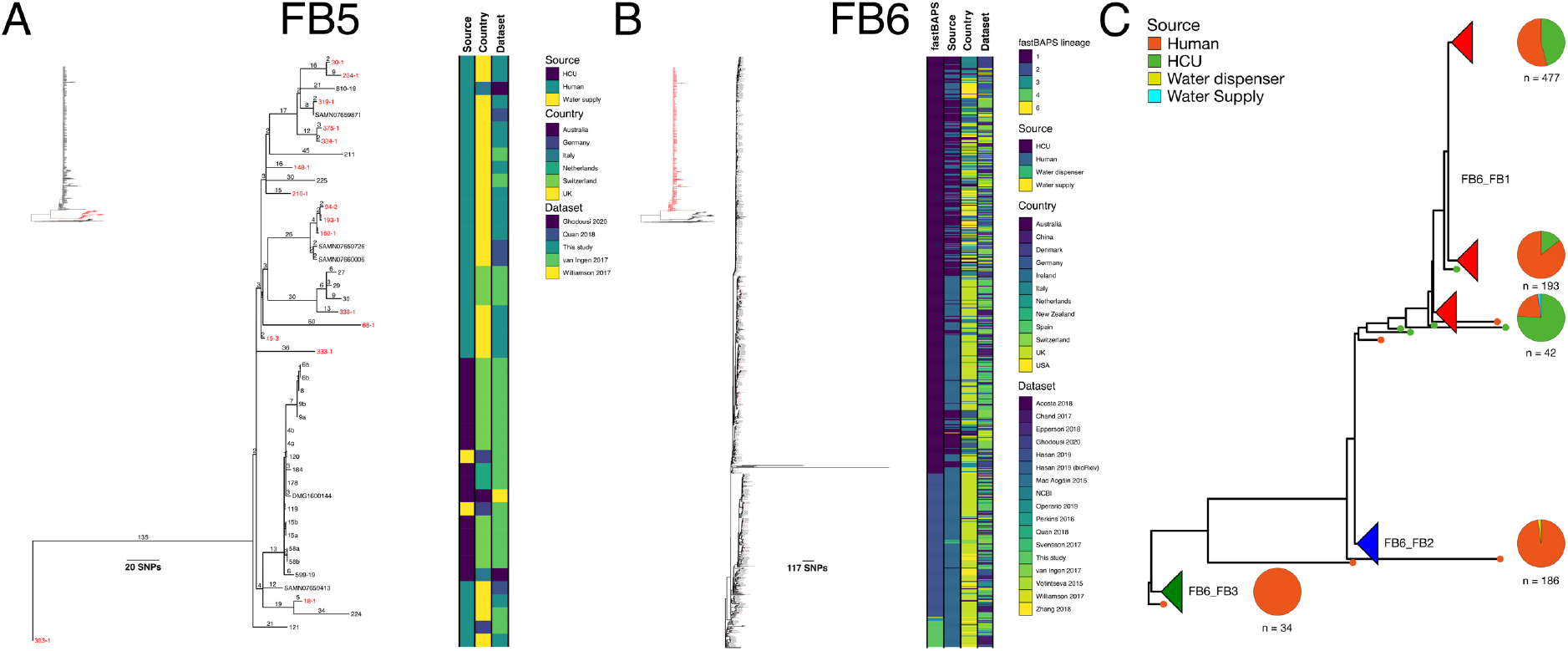
*Mycobacterium chimaera* global fastBAPS cluster phylogenies. Rooted maximum likelihood phylogenetic trees for A) *Mycobacterium chimaera* FB5 (n = 45); B) *Mycobacterium chimaera* FB6 (n = 765) and C) *Mycobacterium chimaera* FB6 with major clades collapsed; pie charts are shown for each major clade showing the distribution of sources for that clade. The taxa are clustered according to their sequence similarities. The lengths of the branches are scaled in nucleotide substitutions per site. The source of isolate, country of collection and study are shown as datastrips to the right of the phylogenies. Isolates from the Royal Brompton Hospital are highlighted in red. Branch lengths are shown in SNPs per site for *Mycobacterium chimaera* FB5.

**Fig 8:**
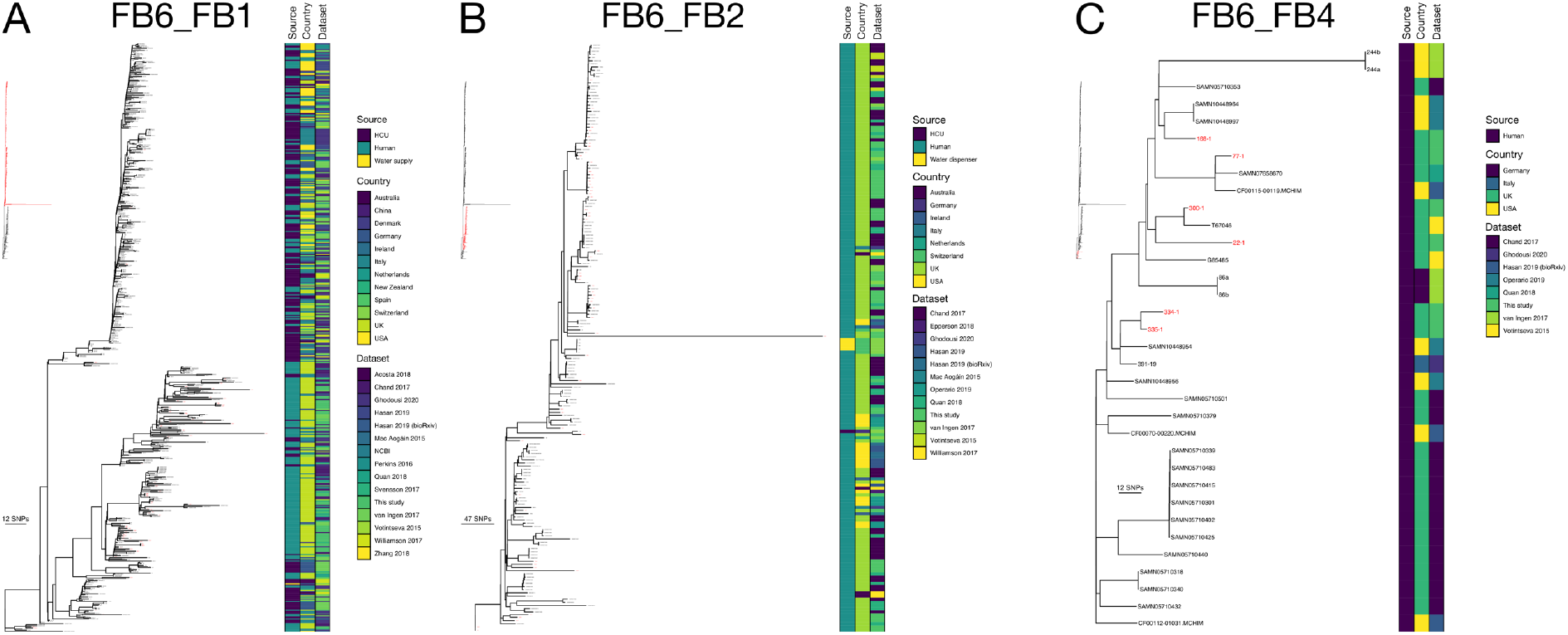
*Mycobacterium chimaera* fastBAPS cluster FB6 phylogenies. Maximum likelihood phylogenetic trees for A) FB6_FB1 (n = 540); B) FB6_FB2 (n = 186); and C) FB6_FB4 (n = 34). The taxa are clustered according to their sequence similarities. The lengths of the branches are scaled in nucleotide substitutions per site. The source of isolate, country of collection and study are shown as datastrips to the right of the phylogenies. Isolates from the Royal Brompton Hospital are highlighted in red.

## Discussion

The aims of this study were to characterise the population structure of MAC and identify potential transmission between CF and non-CF patients. We used a dataset of 996 sequenced MAC isolates collected in a London hospital. This is, to date, the largest sequenced MAC isolate collection analysed from a single location. We showed that polymicrobial infection with MAC is not uncommon. The large, longitudinal dataset enabled us to identify putative transmission clusters for all four species/sub-species we considered. Most of these transmission clusters contained both patients with CF and non-CF patients, showing that transmission of MAC is occurring despite the presence of hospital-based control measures focussing on patients with CF. The lack of clear epidemiological links suggests that the majority of transmission is not due to direct patient-to-patient transmission but rather through intermediates such as a local environmental vector or asymptomatic carriage in the wider population. Comparison of the isolates collected as part of this study with published isolates identified a small number of lineages likely circulating in the UK. With the exception of the major HCU-associated lineage in *M. chimaera*, the comparison did not identify globally circulating lineages like those observed in *M. abscessus*. Surprisingly, given the previous description of the *M. chimaera* HCU clade as representing an environmental point source outbreak (12), we provide evidence that the lineage which was responsible for the HCU-vectored *M. chimaera* outbreak was derived from an existing lineage which was already circulating amongst patients with pre-existing lung conditions, and that this introduction into HCUs happened on at least two separate occasions (only one of which led to the global outbreak). This indicates that, while infection and distribution of HCUs was clearly responsible for the global transmission of one sub-lineage, they were not the primary source of this lineage, which was derived from a lineage that was already circulating in the human population prior to the HCU outbreak. Further, there is evidence of onward transmission into non-cardiac patients of the HCU-sub-lineage, indicating that local establishment and transmission can still occur independent of direct infection by HCUs.

During the isolate sampling period we identified 45 patients infected with more than one species or lineage. This suggests that many of the patients had polymicrobial infections and that certain lineages were preferentially sampled and cultured at different times. This has been observed previously in other longitudinal samples of patients with MAC lung disease (38). These observations suggest that single colony picks during culturing potentially may be underrepresenting the underlying diversity of MAC species in patients with MAC lung disease. Therefore, to better understand the ecology and transmission of MAC species in the lung, a deep sequencing approach in which colony sweeps rather than colony picks are utilised should be considered for future studies.

Previous studies have used SNP thresholds of 15-25 SNPs to identify putative transmission clusters in NTM species (13,18,33). Rather than using a previously applied or arbitrary threshold, we used a method that had previously been used to define SNP thresholds in *Staphylococcus aureus* (55). This allowed us to calculate a SNP threshold for each species using the longitudinal isolates collected by the RBH. The thresholds calculated for *M. intracellulare* and MAH of 16 SNPs were similar to those previously applied. Whilst we obtained higher thresholds of 30 SNPs and 58 SNPs for *M. chimaera* and MAA respectively, this did not result in an inflated number of transmission clusters, as applying a threshold of 16 SNPs to the MAA dataset would have resulted in only two fewer clusters. These larger values were predominantly due to high levels of within-host diversity observed amongst patient isolates in single clusters in both MAA and *M. chimaera*. For instance, in MAA_FB14 isolates from a single patient collected a day apart were as much as 101 SNPs different. This highlighted the likelihood of polymicrobial infections being common amongst the patients we included in the study.

Using the SNP thresholds calculated for each species, we were able to identify putative transmission clusters amongst all four of the MAC species/subspecies we considered. Broadly, two different patterns of infection were observed: amongst *M. intracellulare* and the two *M. avium* subspecies, our results suggested that there have been multiple independent introductions into the human population followed by limited transmission. Our analysis of *M. chimaera*, however, identified a single large transmission cluster was responsible for infecting 106/155 (68.4%) of our patients infected with this species. Most of these transmission clusters contained both CF and non-CF patients and a small number also contained patients with no pre-existing lung conditions.

Using our criteria, we were unable to identify potential epidemiological links for most patients included in the transmission clusters. The exception to this was the largest *M. chimaera* transmission cluster where we found 12 potential epidemiological links up to a year before sampling began. Interestingly, four patients were part of more than one link, with patient 218 overlapping with other patients in this transmission cluster on the same wards six times over a nine-month period. The tight infection control surrounding patients with CF, which prioritises preventing cross-infection through hygiene and segregation, means that they are unlikely to have interacted with patients with other lung conditions whilst in hospital. Outpatient clinics are organised so that patients are seen individually and, during inpatient care, CF patients are given individual rooms with en-suite facilities. This implies that transmission is likely occurring *via* other pathways, such as through environmental intermediates or through a reservoir of healthy, asymptomatic carriers in the wider population. This observation is not unique and for some time, there has been considerable debate whether NTM transmission between patients is occurring, even when transmission is strongly supported by genome-based analysis (13,18,58). The near complete absence of epidemiological links between patients in transmission clusters as well as the ubiquitous presence of NTM species in the environment, especially water supplies, has led some researchers to suggest that the dissemination of NTM lineages at a national level is associated with exposure to contaminated water supplies (18). Whilst there is certainly strong evidence for the same lineages being isolated from water supplies and patients in the same location [28], the absence of a single national water supply even in a comparatively small country like the UK would suggest that closely related isolates collected from different geographical locations are unlikely to be due to a single contaminated water supply. It is however possible that transmission networks may include local water supplies as intermediates. The alternative hypothesis that NTM populations are being maintained in apparently healthy individuals with no symptoms of NTM lung disease is equally worthy of consideration. This would explain the presence of local transmission clusters in the absence of direct epidemiological links as well as the presence of phylogeographical structure in NTM trees. To better address how NTM species are being transmitted, future studies should focus on attempting to collect isolates from the general population, perhaps focussing on smokers where there is some evidence that they may provide a better reservoir for NTM species and other opportunistic pathogens (19).

The inclusion of published genomes alongside the RBH isolates allowed us to use our defined pairwise SNP thresholds to identify putative MAC lineages circulating in the UK and globally. Apart from the *M. chimaera* lineages that were known to be associated with HCUs, we were unable to identify large globally circulating lineages like those observed in *M. abscessus*. This potentially suggests different evolutionary dynamics amongst the MAC species or, in the case of *M. intracellulare*, may simply reflect a lack of comprehensive sampling. We were only able to identify a single lineage containing RBH isolates amongst the two *M. avium* subspecies that included isolates from outside the UK. This cluster comprised the majority of isolates in FB15 (Figure 5B). Given that most available *M. avium* genomes have been collected in the UK or USA, a more comprehensive global sampling strategy could potentially reveal the existence of additional globally circulating clones.

A different picture emerged when we calculated global transmission clusters for *M. chimaera* using a pairwise SNP threshold of 30 SNPs (Supplementary Table 7). Here we found that most clusters either consisted of isolates from across Europe or else also included isolates from other parts of the world such as Australia. Notably, we identified a single large transmission cluster containing 489 isolates from 12 countries that included most of the HCU-associated isolates. Previous work investigating the global population structure of *M. chimaera* has suggested that the majority of *M. chimaera* transmission globally was associated with point source contamination of HCUs during manufacturing which then led to onward transmission to patients undergoing cardiac surgery (12). In our study we have assembled the largest *M. chimaera* dataset analysed to date, including isolates collected from HCUs and cardiac patients as well as from patients with lung conditions such as CF and BX. This has enabled us to investigate the global population structure of *M. chimaera* in more detail than has previously been possible (12,59). Strikingly, we discovered clear evidence for distinct lineages within the global phylogeny that were almost exclusively associated with patient isolates. In addition, most of the HCU-associated isolates were clustered together in a single lineage that was derived from the lineages containing only human isolates (Figure 6B). This was also true of a second, smaller, lineage that contained only HCU and environmental isolates. The results suggest that the predominant HCU-associated lineage is descended from a *M. chimaera* population already circulating amongst patients with respiratory diseases such as CF, and that a similar derivation has happened at least once more. Unfortunately, due to the absence of even a moderate temporal signal (data not shown), we were unable to place any estimates of when these lineages first emerged.

Potential limitations of this study are the lack of environmental samples as well as the distinct bias with respect to country of isolation in the contextual dataset. Given, the lack of direct patient to patient contact and epidemiological links, the inclusion of samples collected from the wards admitting patients and hospital water supplies would potentially have revealed additional vectors of transmission. Assembling useful contextual datasets is reliant on what is available in the public domain and the geographical bias towards the USA and UK in the contextual datasets we used is not confined to this study. For many pathogens, most studies have focussed on resource richer settings. However, given the increasing availability of genome sequencing, it is likely that NTM datasets from other settings will be available to include in future analyses.

In conclusion, using a large longitudinal sample of MAC isolates collected within a single hospital as well as previously published global contextual isolates, we were able to identify polymicrobial infections by different MAC species and lineages in multiple patients as well as identify transmission clusters representing lineages circulating within the hospital, at a national level and, less commonly, globally. We show that the origin of the major HCU-associated *M. chimaera* lineage is likely a previously circulating lineage associated with patients with respiratory diseases. To better understand the transmission dynamics and to prevent continuing circulation of MAC and other NTM species, future studies should include sampling of environmental reservoirs and potential asymptomatic carriers.

## Data Availability

Raw sequencing reads were deposited at the European Nucleotide Archive under project PRJEB21813; all accessions used in this project are listed in Supplementary File 1.

## Code availability

The R code used to perform data analyses in this study is available in GitHub (https://github.com/avantonder/MAC).

## Supplementary Material

**Table A.**
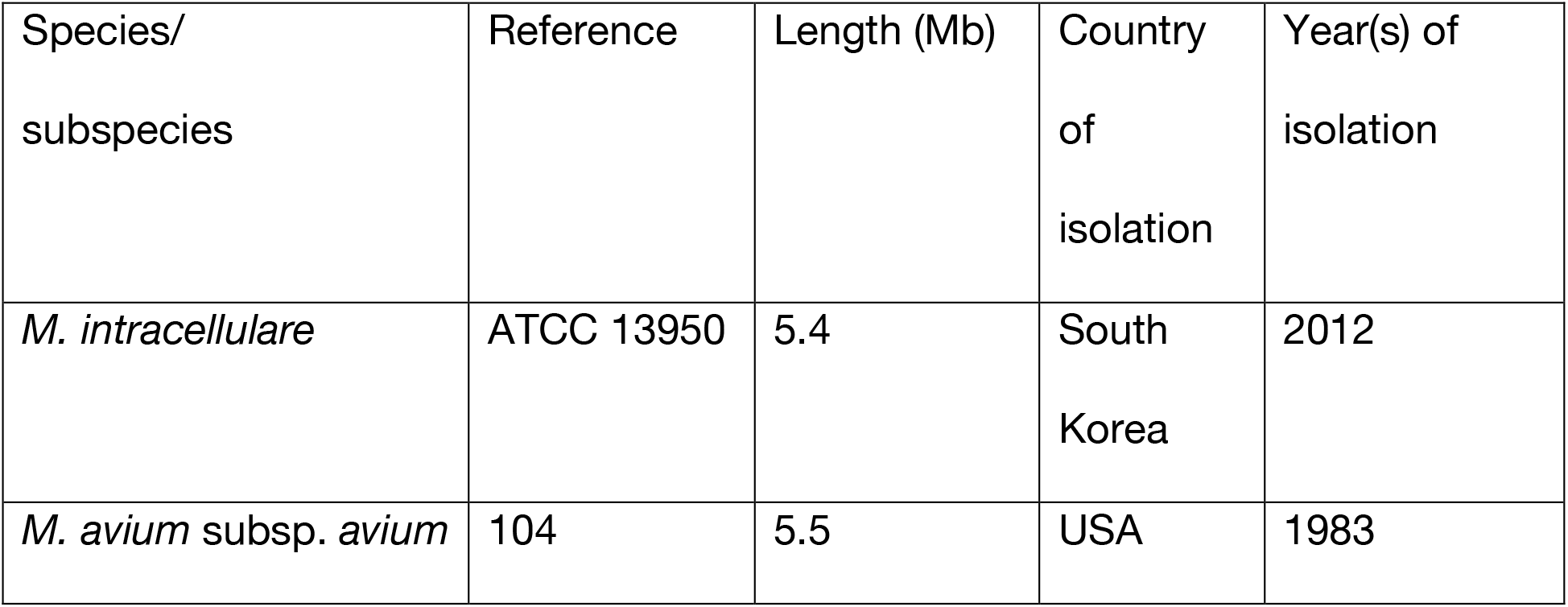
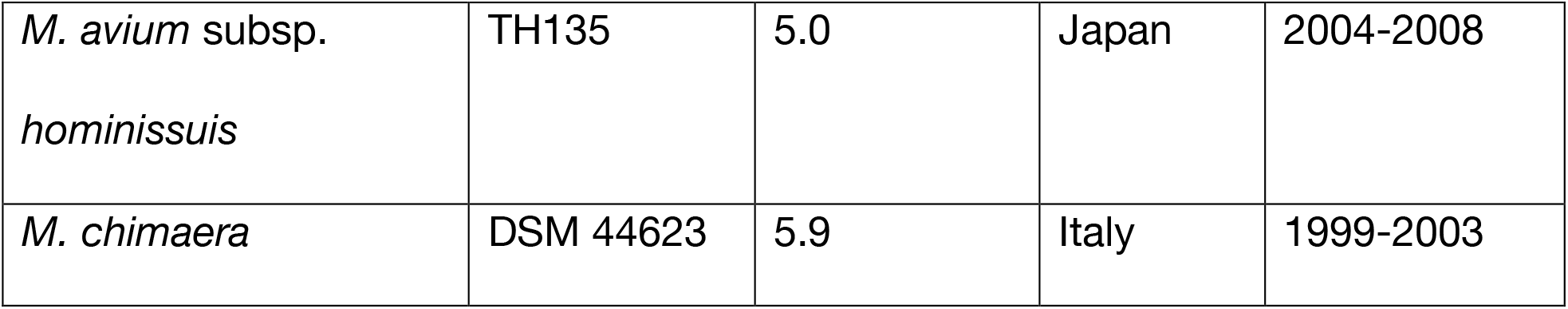
Reference genomes used in this study.

**Table B.** *Mycobacterium intracellulare* transmission clusters

| fastBA<br>PS<br>cluster | Transmiss<br>ion cluster | Patien<br>ts (n) | Bronchiect<br>asis (n) | Cystic<br>Fibros<br>is (n) | Other<br>lung<br>conditio<br>ns (n) | No<br>pre-<br>existin<br>g lung<br>conditi<br>on (n) | Patients<br>undergoi<br>ng<br>treatmen<br>t (n) |
| --- | --- | --- | --- | --- | --- | --- | --- |
| Mi_FB2 | Mi_FB2_1 | 2 | 0 | 2 | 0 | 0 | 1 |
| Mi_FB3 | Mi_FB3_1 | 16 | 8 | 7 | 1 | 0 | 4 |
| Mi_FB5 | Mi_FB5_1 | 3 | 2 | 0 | 1 | 0 | 2 |

**Table C.** *M. avium* subsp. *avium* transmission clusters

| fastBAP<br>S cluster | Transmiss<br>ion cluster | Patien<br>ts (n) | Bronchiect<br>asis (n) | Cysti<br>c<br>Fibro<br>sis (n) | Other<br>lung<br>conditio<br>ns (n) | No<br>pre-<br>existin<br>g lung<br>conditi<br>on (n) | Patients<br>undergoi<br>ng<br>treatme<br>nt (n) |
| --- | --- | --- | --- | --- | --- | --- | --- |
| MAA_FB<br>5 | MAA_FB5<br>_1 | 10 | 6 | 2 | 2 | 0 | 4 |
| MAA_FB<br>5 | MAA_FB5<br>_4 | 2 | 1 | 1 | 0 | 0 | 0 |
| MAA_FB<br>5 | MAA_FB5<br>_5 | 2 | 1 | 0 | 1 | 0 | 1 |
| MAA_FB<br>5 | MAA_FB5<br>_12 | 2 | 1 | 1 | 0 | 0 | 1 |
| MAA_FB<br>7 | MAA_FB7<br>_1 | 2 | 2 | 0 | 0 | 0 | 1 |
| MAA_FB<br>10 | MAA_FB1<br>0_1 | 2 | 0 | 2 | 0 | 0 | 0 |
| MAA_FB<br>14 | MAA_FB1<br>4_1 | 2 | 2 | 0 | 0 | 0 | 1 |

**Table D.** *M. avium* subsp. *hominissuis* transmission clusters

| fastBAP<br>S cluster | Transmissi<br>on cluster | Patie<br>nts (n) | Bronchiect<br>asis (n) | Cysti<br>c<br>Fibro<br>sis (n) | Other<br>lung<br>conditi<br>ons (n) | No<br>pre-<br>existin<br>g lung<br>conditi<br>on (n) | Patients<br>undergo<br>ing<br>treatme<br>nt (n) |
| --- | --- | --- | --- | --- | --- | --- | --- |
| MAH_F<br>B8 | MAH_FB8<br>_1 | 16 | 7 | 4 | 3 | 1 | 0 |
| MAH_F<br>B10 | MAH_FB1<br>0_1 | 4 | 2 | 2 | 0 | 0 | 1 |
| MAH_F<br>B11 | MAH_FB1<br>1_1 | 6 | 3 | 1 | 2 | 0 | 0 |
| MAH_F<br>B12 | MAH_FB1<br>2_2 | 4 | 1 | 2 | 1 | 0 | 2 |
| MAH_F<br>B14 | MAH_FB1<br>4_3 | 4 | 2 | 2 | 0 | 0 | 0 |
| MAH_F<br>B14 | MAH_FB1<br>4_7 | 2 | 0 | 2 | 0 | 0 | 0 |
| MAH_F<br>B14 | MAH_FB1<br>4_9* | 7 | 3 | 2 | 1 | 0 | 1 |

**Table E.** *Mycobacterium chimaera* transmission clusters

| fastBA<br>PS<br>cluster | Transmiss<br>ion cluster | Patien<br>ts (n) | Bronchiect<br>asis (n) | Cystic<br>Fibros<br>is (n) | Other<br>lung<br>conditio<br>ns (n) | No<br>pre-<br>existin<br>g lung<br>conditi<br>on (n) | Patients<br>undergoi<br>ng<br>treatmen<br>t (n) |
| --- | --- | --- | --- | --- | --- | --- | --- |
| Mc_FB<br>3 | Mc_FB3_<br>1 | 106 | 43 | 24 | 25 | 3 | 15 |
| Mc_FB<br>3 | Mc_FB3_<br>3 | 2 | 1 | 1 | 0 | 0 | 0 |
| Mc_FB<br>3 | Mc_FB3_<br>4 | 2 | 2 | 0 | 0 | 0 | 0 |
| Mc_FB<br>3 | Mc_FB3_<br>6 | 2 | 1 | 0 | 1 | 0 | 0 |
| Mc_FB<br>3 | Mc_FB3_<br>13 | 4 | 1 | 1 | 1 | 1 | 1 |
| Mc_FB<br>3 | Mc_FB3_<br>16 | 2 | 0 | 0 | 1 | 1 | 1 |
| Mc_FB<br>3 | Mc_FB3_<br>17 | 2 | 1 | 1 | 0 | 0 | 0 |
| Mc_FB<br>3 | Mc_FB3_<br>19 | 2 | 0 | 1 | 0 | 1 | 0 |
| Mc_FB<br>3 | Mc_FB3_<br>25 | 2 | 1 | 1 | 0 | 0 | 1 |
| Mc_FB<br>3 | Mc_FB3_<br>30* | 4 | 0 | 1 | 0 | 0 | 0 |
| Mc_FB<br>4 | Mc_FB4_<br>1 | 2 | 1 | 1 | 0 | 0 | 1 |
| Mc_FB<br>4 | Mc_FB4_<br>2 | 9 | 3 | 3 | 3 | 0 | 2 |
| Mc_FB<br>4 | Mc_FB4_<br>3* | 6 | 1 | 2 | 2 | 0 | 1 |

**Table F.** *Mycobacterium chimaera* epidemiological links

| Patients with epidemiological link | Hospital ward | Dates of overlapping stay |
| --- | --- | --- |
| 24/218 | FOUL | 02/02/12-12/02/12 |
| 163/218 | FOUL | 13/02/12 |
| 175/218 | FOUL | 05/04/12-16/04/12 |
| 130/218 | LIND | 30/04/12 |
| 76/218 | FOUL | 06/06/12-11/06/12 |
| 32/163 | LIND | 29/06/12 |
| 79/241 | LIND | 05/07/12 |
| 241/272 | LIND | 06/07/12 |
| 177/186 | LIND | 06/08/12 |
| 122/175/306 | FOUL | 25/10/12-30/10/12 |
| 30/122 | FOUL | 05/11/12-06/11/12 |
| 30/218 | FOUL | 12/11/12-13/11/12 |

**Table G.** *Mycobacterium chimaera* global transmission clusters containing RBH isolates

| fastBAPS cluster | Transmission cluster | Total isolates (n) | Patients (n) | HCU (n) | Other (n) |
| --- | --- | --- | --- | --- | --- |
| FB5 | FB5_1 | 21 | 4 | 15 | 2 |
| FB5 | FB5_2 | 7 | 7 | 0 | 0 |
| FB5 | FB5_3 | 5 | 5 | 0 | 0 |
| FB5 | FB5_6 | 4 | 4 | 0 | 0 |
| FB6_FB1 | FB6_FB1_1 | 489 | 230 | 258 | 1 |
| FB6_FB1 | FB6_FB1_2 | 6 | 6 | 0 | 0 |
| FB6_FB1 | FB6_FB1_8 | 5 | 5 | 0 | 0 |
| FB6_FB1 | FB6_FB1_21 | 4 | 4 | 0 | 0 |
| FB6_FB2 | FB6_FB2_1 | 22 | 22 | 0 | 0 |
| FB6_FB2 | FB6_FB2_2 | 36 | 36 | 0 | 0 |
| FB6_FB2 | FB6_FB2_8 | 16 | 16 | 0 | 0 |
| FB6_FB2 | FB6_FB2_17 | 10 | 10 | 0 | 0 |
| FB6_FB2 | FB6_FB2_32 | 11 | 11 | 0 | 0 |
| FB6_FB2 | FB6_FB2_35 | 2 | 2 | 0 | 0 |
| FB6_FB4 | FB6_FB4_8 | 4 | 4 | 0 | 0 |
| FB6_FB4 | FB6_FB4_11 | 3 | 3 | 0 | 0 |
| FB6_FB4 | FB6_FB4_16 | 2 | 2 | 0 | 0 |

**Fig A.**
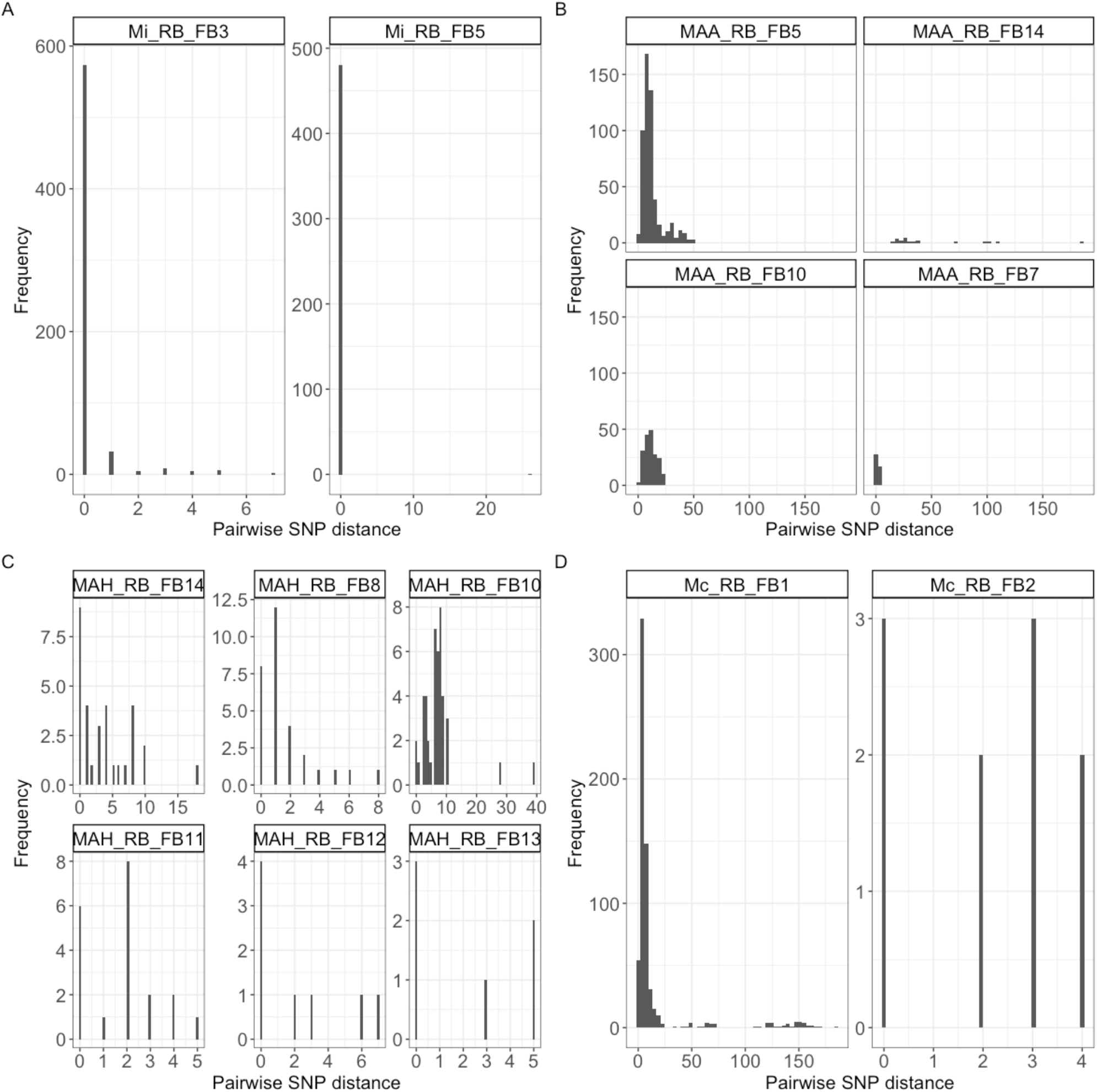
Within-host isolate pairwise SNP diversity in fastBAPS lineages for A) *Mycobacterium intracellulare*; B) *Mycobacterium avium* subsp. *avium;* C) *Mycobacterium avium* subsp. *hominissuis* and D) *Mycobacterium chimaera*.

**Fig B.**
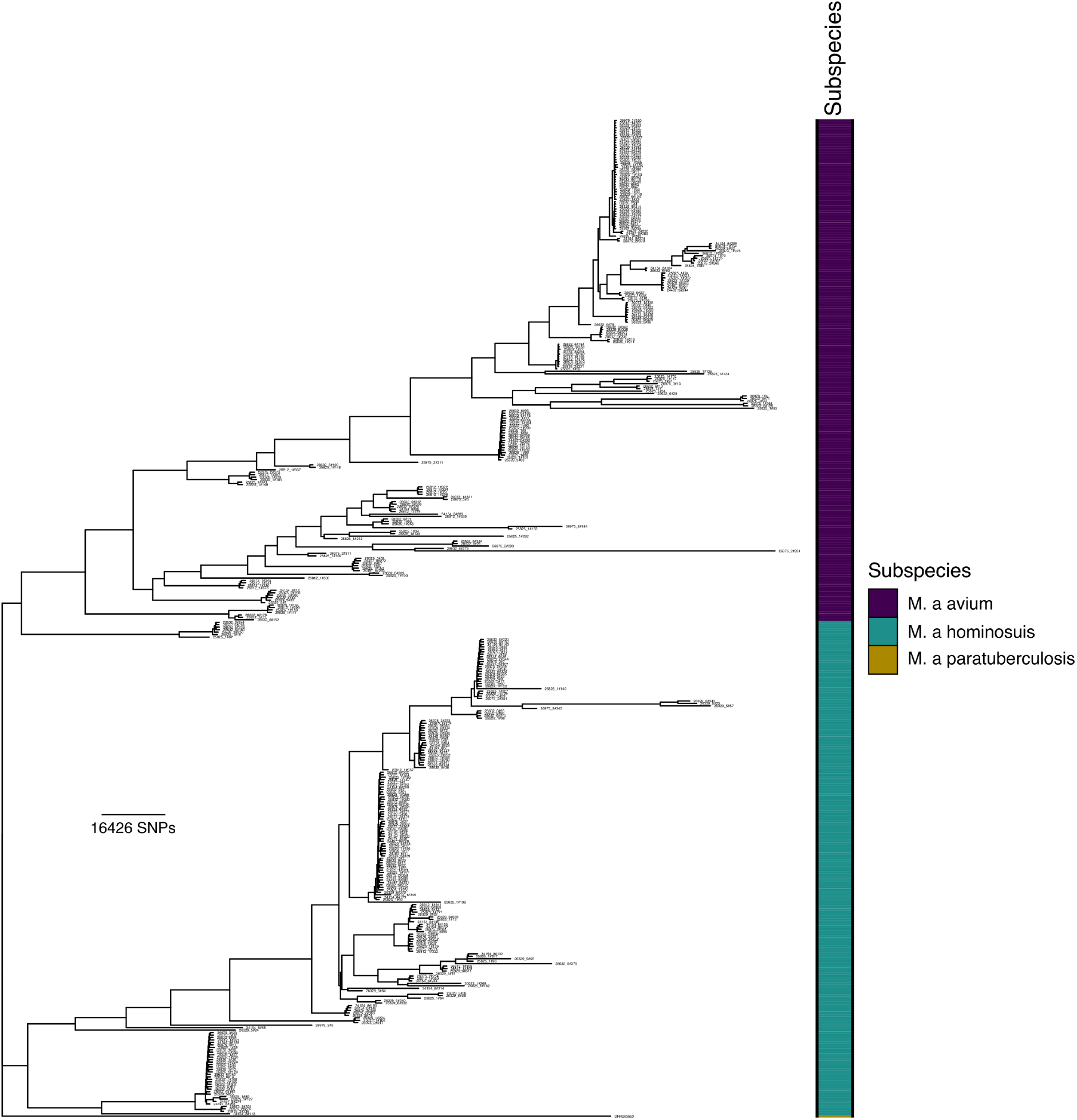
Population structure of *Mycobacterium avium* at the Royal Brompton Hospital. Maximum likelihood phylogenetic tree of 406 *Mycobacterium avium* isolates rooted with a *Mycobacterium avium* subsp. *paratuberculosis* isolate (DRR263663). The subspecies of each isolate is shown in the datastrip to the right of the phylogeny. The scale bar is shown in SNPs per site.

**Fig C.**
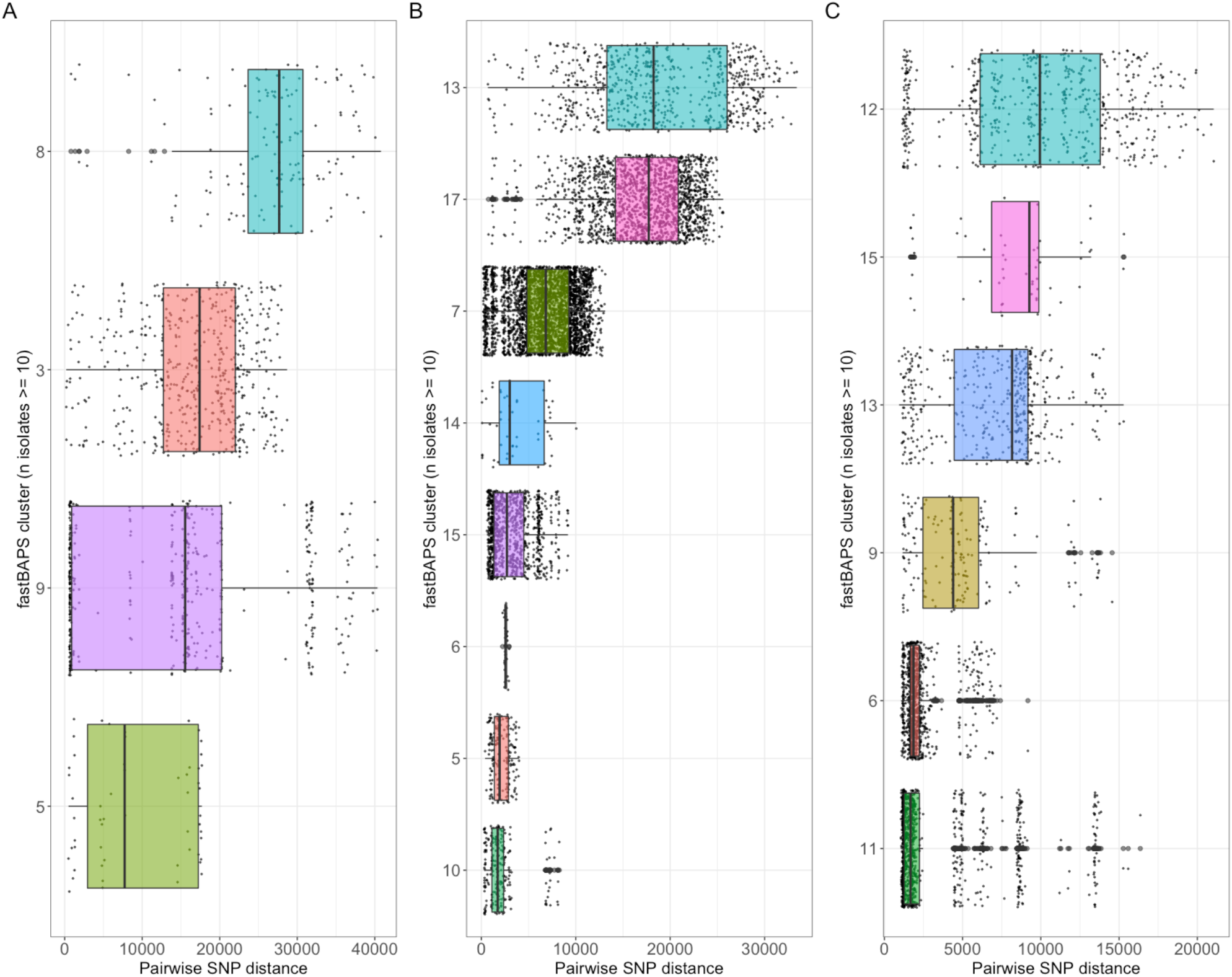
Global fastBAPS cluster pairwise SNP distance distributions. Boxplots showing isolate pairwise SNP distances for A) *Mycobacterium intracellulare*; B) *Mycobacterium avium* subsp. *avium* and C) *Mycobacterium avium* subsp. *hominissuis*

**Fig D:**
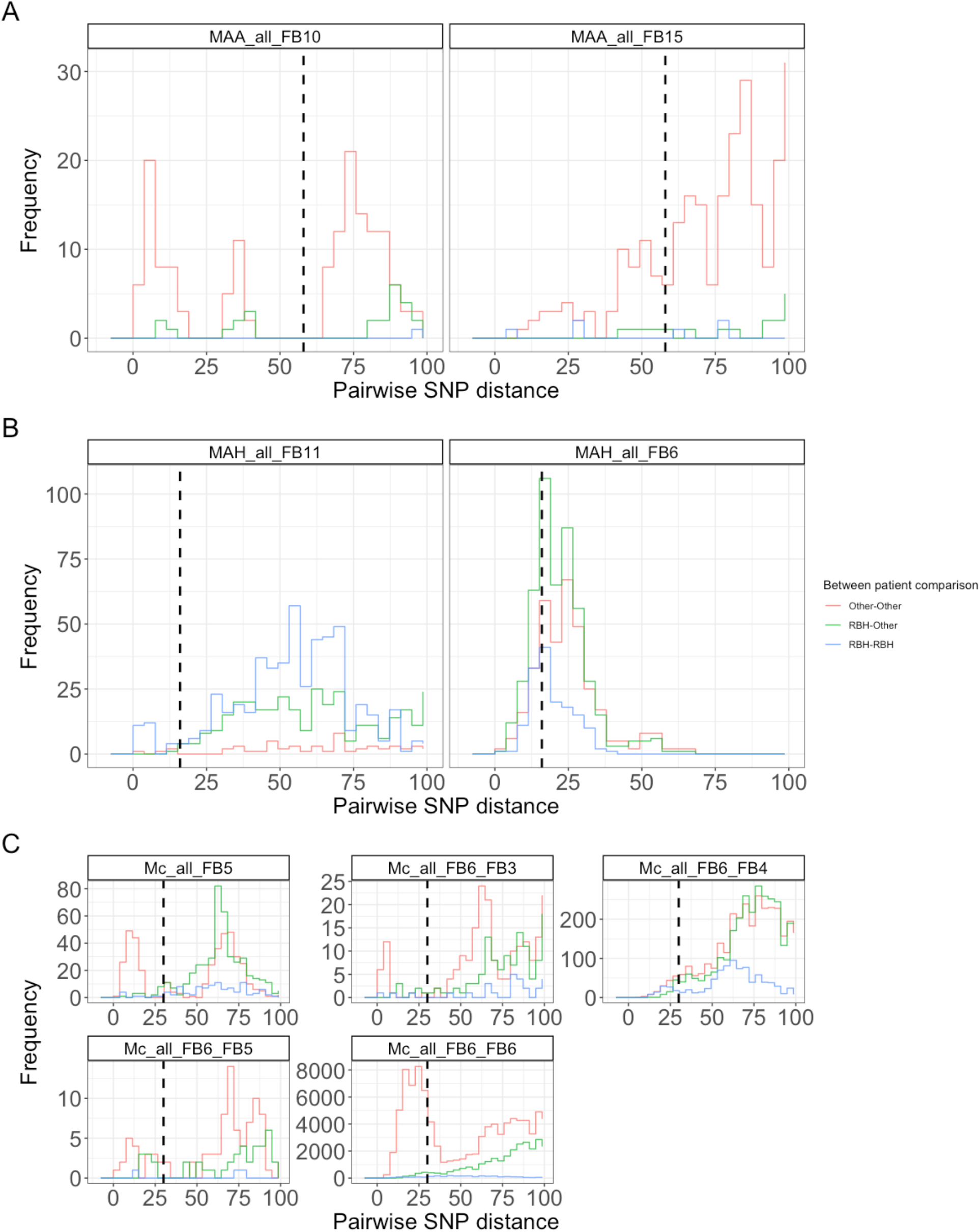
Distribution of pairwise SNP distances between isolates from the Royal Brompton Hospital and other studies. Histograms showing distribution of pairwise SNP distances for fastBAPS clusters for A) *Mycobacterium avium* subsp. *avium*; B) *Mycobacterium avium* subsp. *hominissuis* and C) *Mycobacterium chimaera*.

